# Understanding the Impact of Heatwaves on UK Care Homes: A National Survey of Staff Experiences, Challenges, and Adaptation Strategies

**DOI:** 10.64898/2026.03.24.26349157

**Authors:** Hannah Blount, Jade Ward, Patrick AB James, Peter R Worsley, Davide Filingeri, Nuno Koch Esteves

**Affiliations:** ThermosenseLab, Skin Sensing Research Group, School of Health Sciences, University of Southampton, Southampton, UK; Tyndall Centre for Climate Change Research, University of Southampton, Southampton, UK; School of Engineering, Faculty of Engineering and Physical Sciences, University of Southampton, Southampton, UK; PressureLab, Skin Sensing Research Group, School of Health Sciences, University of Southampton, Southampton, UK

**Author notes:** Joint first authors. Co-first author HB and co-first author JW, contributed equally to this work. **Corresponding author:** <u>Dr Nuno Koch Esteves</u> ThermosenseLab Skin Sensing Research Group (SSRG) Clinical Academic Facility | Level A, Room AA83 | South Academic Block (MP11) Southampton General Hospital | Tremona Road | SO16 6YD School of Health Science, University of Southampton, UK.

**Keywords:** Health Personnel, Environmental Medicine, Environmental Monitoring, Occupational Medicine, Public Health

## Abstract

**Introduction:** Climate change is increasing the frequency and intensity of heatwaves, creating critical challenges for social care settings where both staff and residents face heightened heat⍰related vulnerability. This study examined the impact of heatwaves on UK care homes using a national survey of staff experiences, challenges, and adaptation strategies.

**Methods:** Care home staff (*N* = 225) in managerial (*N* = 88) and caregiving roles (*N* = 137) completed an online survey investigating staff perceptions of heatwaves impact on thermal comfort, health and vulnerability of themselves and residents, alongside current heat resilience strategies and the barriers to their implementation.

**Results:** Two thirds (66%) of the surveyed staff complained of being too hot three or more times per day resulting in a perceived impact on their ability to perform tasks (90%) and on residents’ comfort and health (92%). Staff demonstrated strong awareness of older adults’ heightened heat vulnerability (95%) and signs of heat illness (87%). Thematic analysis identified five key barriers to providing effective cooling: funding limitations, inadequate equipment, building constraints, staffing pressures, and individual resident needs; and four priority improvement areas: increased access to cooling equipment, improved temperature control, strengthened strategy and policy, and support for staff needs.

**Conclusions:** Heatwaves place considerable strain on care homes, challenging staff capacity to maintain comfortable thermal conditions, despite good knowledge of heat risks. Financial, infrastructural, and staffing constraints limit effective heat⍰resilience practices. Evaluating and implementing affordable, accessible, and context⍰appropriate cooling strategies will be essential to protect both residents and staff as extreme heat events become more frequent.

**KEY MESSAGES:** *WHAT IS ALREADY KNOWN ON THIS TOPIC:*

- Older adults in care homes experience elevated mortality during heatwaves, and these events increase the workload and strain on care staff.
- Most social care practitioners demonstrate a reactive approach when responding to extreme high temperatures.

*WHAT THIS STUDY ADDS:*

- This study provides UK⍰wide evidence that care home staff possess strong knowledge of heat risks, resident vulnerability, and how to recognise signs of heat illness.
- It reveals that despite this knowledge, staff face substantial barriers—most notably inadequate cooling equipment, restricted ventilation, ageing buildings, and heavy workloads—that prevent effective heat⍰resilience practice.
- Care home staff clearly identified the sector’s priority challenges: limited funding, insufficient or ineffective cooling systems, building constraints, staffing pressures, and the diverse thermal needs of residents.

*HOW THIS STUDY MIGHT AFFECT RESEARCH, PRACTICE OR POLICY:*

- Findings demonstrate that current heat⍰resilience measures in UK care homes are insufficient, with staff reporting routine thermal discomfort and compromised working conditions during heatwaves. Evaluating and implementing low⍰cost, effective cooling strategies, and embedding them into policy and practice, will be essential to safeguard both residents and staff as extreme heat events become more frequent.

## 1. Introduction

Global average surface temperature projections estimated by the Met Office (1) predict a likely rise in temperatures of up to 5.4°C by 2070. As a result, the intensity and frequency of extreme weather events, including heatwaves, are likely to increase globally (2), including in the UK (1). Heatwaves already represent the leading cause of weather-related mortality worldwide (3), and they pose severe health risks to vulnerable groups, notably older adults and those with long-term conditions (3). For example, the 2022 European heatwave resulted in an increased mortality rate between 14% and 27% in people over the age of 65 (4). Alarmingly, during heatwaves, mortality increases by ∼10% among older adults residing in care homes (also called residential aged care homes) (5). Hence, the prevention of these heat-related deaths is an increasing public health priority (6, 7).

Ageing is known to impair one’s ability to regulate body temperature, heightening vulnerability to heat stress through attenuations in cardiac output and skin vasodilation, and a diminished sweating responses (8–10). Furthermore, ageing can decrease one’s thermal sensitivity and the resulting ability to behaviourally adapt to heat stress (11). Whilst the impact of heatwaves on the health of older adults has been frequently documented, especially the association between high temperatures, mortality and morbidity (12–14), very few studies have investigated the heat resilience of social care settings such as care homes (15, 16). The UK Health Security Agency (UKHSA) recently investigated the experience of social care practitioners in relation to extreme temperatures (17), and reported that most social care practitioners tend to take a reactive approach when responding to extreme high temperatures. However, this research did not investigate whether barriers to the implementation of proactive heat resilience strategies exist within social care settings. This knowledge gap is particularly relevant for UK care settings, which are characterised by increasing heat exposure, yet limited structural and organisational preparedness (18). Importantly, rising indoor temperatures not only endanger residents but also increase the physical and cognitive demands placed on care home staff, whose workload intensifies during heatwaves as they support residents’ thermal safety (19, 20). This additional burden may render staff themselves increasingly heat-vulnerable, particularly when working in inadequately cooled environments.

Heat resilience consists of proactively managing and mitigating both acute and chronic extreme heat exposures (21). In this respect, the UKHSA advises that indoor temperatures in care homes should not exceed 26°C to protect vulnerable residents (17). However, recent field data on indoor ambient temperatures from care homes in London (UK) revealed summertime temperatures exceeding 30°C during day-time hours in heatwave periods (22). These observations highlight ongoing risks for care home residents and care staff, who may experience intensified workload and occupational heat exposure, whilst working in conditions exceeding UKHSA guidelines, thereby contributing to heat-related negative health outcomes.

Owing to residents’ care needs and physiological states—e.g., reduced autonomic and behavioural capacity to withstand heat (23)—the role of care home staff is vital in ensuring appropriate heat management and thermal protection during heatwave periods. Despite the recognised importance of care home staff in maintaining safe thermal environments, their ability to implement effective heat management practices may be constrained by several factors, including limited training, competing care priorities, infrastructural limitations, and insufficient organisational guidance (24–27). Better understanding these barriers from a staff’s perspective, alongside the strategies that staff employ to protect residents during extreme heat, is essential to improve the heat resilience of social care settings and their residents.

The aim of this study was three-fold: (*i*) to understand the impact of heatwaves on care home staff’s, and their perceptions of residents’, health and comfort; (*ii*) to evaluate the heat resilience strategies employed by care home staff during heatwave periods; and (*iii*) to explore the barriers to their implementation. To achieve these aims, a survey was design and deployed across the UK and Ireland to capture care home staff’s experiences, challenges, and adaptation strategies during heatwaves. Our overarching goal was to use the survey data to identify patterns in heat (mal-)adaptation that could be used to inform improved, evidence-based heat-resilience policies within social care settings.

## 2. Materials and Methods

### 2.1 Ethical Approval

This study was approved by the University of Southampton Ethics Committee (approval No.103283), and all procedures conformed to the ethical standards set by the Declaration of Helsinki. Participants were informed that this was a voluntary survey, and written informed consent was obtained prior to the survey completion. Monetary incentives in the form of the opportunity to win one of forty £50 Amazon vouchers were available. Data collected were stored on a secure database at the University of Southampton, and only the research team could access the anonymised survey results.

### 2.2 Participants

A convenience sampling approach was used to recruit care home staff in both managerial (e.g., manager, deputy manager, team leader, administration) and care giver (e.g., nurse, care worker, senior care worker, personal resident care assistant) roles. These roles were selected to accommodate an understanding of both administrative perspectives on heat management within the care setting, as well as care delivery perspectives from care. Care staff are likely to have the most resident contact, thereby providing representative insights on care delivery and residents’ vulnerability during periods of extreme hot weather.

Over 300 private and public care homes were contacted from the UK and Ireland. Eligible participants had to be employed by such organisation and meet the following Inclusion and exclusion criteria are listed below:

#### Inclusion

- Aged 18 and over.
- Employed staff member working in an older adult care home, i.e., not agency staff
- Holds either a managerial or care giver role.

#### Exclusion

- Unable to complete the survey in English.
- Residents of the care home.
- Non-care giver or managerial roles (i.e. maintenance or kitchen staff)

The umbrella term for *care homes* can encompass different definitions (28). Data was collected from individuals who worked in each of the different care home types. However, as the term *care home* is discussed across the current study, it refers to the overall setting rather than a specific type of care:

- Care home: helps with personal care such as washing, dressing, taking medication and going to the toilet;
- Nursing home: offers personal care as well as 24-hour assistance from qualified nurses;
- Dual registered care home: accepts residents who need both personal care and nursing care, i.e., if someone’s needs increase over time, they would not have to move to a different home.

### 2.3 Data Collection Procedures

A literature review was conducted to establish question styles, questions of interest and the overarching four key sections. Furthermore, various questions were workshopped and adapted using examples and evidence provided in previous survey papers investigating topic areas in care homes, such as thermal comfort (23, 29–32), climate resilience (33), facility preparedness (34, 35), vulnerability (15, 36), thermal perception (22, 37), and heat waves strategies (17, 38, 39).

Prior to data collection, a pilot survey was conducted with care home staff (*N* = 4) to assess question clarity, language, and survey length. Feedback indicated that the survey was generally easy to use but could be improved by adding brief explanations of key terms and reducing completion time. An exploratory Principal Component Analysis (PCA) was carried out to help identify any obvious cross⍰correlations or redundancies among items; however, given the small pilot sample size, the PCA was used only as a preliminary guide. Final decisions regarding item removal were based primarily on participant feedback and the research team’s evaluation of item clarity and relevance, resulting in the refinement of the survey from 48 to 41 questions.

Data was collected from March to September 2025. The online survey (Qualtrics XM 2023, United States) was distributed to care home staff through two main channels: (*i*) direct contact to care home owners and managers (i.e., acting gatekeepers) requesting permission to distribute amongst their staff, or (*ii*) through social media campaigns. Managers and owners of the care homes were contacted directly via an introductory email with a brief description of the survey and study outcomes. Once they agreed to take part, a further email was sent with a URL link and QR code to the survey, the recruitment poster, along with recommendations on how to disseminate/distribute amongst their staff. In addition, the survey was advertised through our THERMOSENSELAB-associated social media platforms. Community groups, organisations and local councils were contacted directly—acting as gatekeepers—who distributed the survey to their network via email, social media and newsletters.

### 2.4 Online questionnaire

The 41-question anonymised survey (*available in supplementary material*) took approximately ten minutes to complete and comprised of multiple-choice and open-ended questions. The survey was separated into four sections: demographics, staff’s and staff’s perceptions of residents’ thermal comfort, heat resilience strategies (knowledge and implementation), and open-ended questions regarding the barriers that exist to current heat resilience strategies. The purpose of these sections was firstly to determine the impact of variations in indoor ambient temperature on care home staff’s health, comfort, and vulnerability, and their perceptions of residents’ health, comfort, and vulnerability to heat-related complications during heatwave periods. Secondly, the survey aimed to gain an overview of the thermal management strategies employed in care homes, alongside an assessment of the strategies’ implementation and associated barriers.

#### 2.4.1 Demographic

This section consisted of 12 items in written or tick⍰box format. This captured key contextual information—such as participants’ age, job role, length of time in their current position, type of care home, and the approximate number of residents—while avoiding any personally identifying details. All questions were designed to ensure that data remained fully anonymous and could not be linked back to individual respondents.

#### 2.4.2 Staff’s and residents’ thermal comfort

This subcategory comprised of ten questions assessed the staff’s perception of their own, and the residents’ thermal comfort, the importance of thermal comfort, and the impact that extreme hot weather has on their satisfaction with their working environment and resident health (31, 37). Most questions comprised of a 1-to-5 point Likert scale, e.g., “on average, how often do staff complain about being too hot?” where 1 is ‘Never’ and 5 is ‘Very often (4+ times per day)’. An additional multiple-choice question was incorporated to indicate the source of dissatisfaction (if any) with the temperature in the workspace.

#### 2.4.3 Heat resilience strategies—knowledge and implementation

This section of eighteen questions aimed to understand the staff’s current knowledge on extreme hot weather such as awareness on guidance, training provided and how capable are the residents at managing their own thermal comfort (34, 35). This section aimed to understand the staff’s ability to manage and assess the thermal comfort of the residents and the care home. In this section, questions comprised of 1-to-5 point Likert scales, multiple choice questions, and a randomised number order rank list. The types of questions asked were, for example, “how regularly do you check the indoor temperature?”, “what is the central heating set to in the summer?”, “who has access to control the indoor temperature?”, and “what cooling strategies are in place?”.

#### 2.4.4 Open-ended questions regarding the barriers that exist to current heat resilience strategies

Finally, participants had the option to complete two open-ended questions. The two questions gave participants the opportunity to express what barriers exist currently in their care home to maintaining thermal comfort for themselves and residents during heatwaves, as well as what improvements or changes they would make to the care home strategy to better manage thermal comfort.

All survey items, except the open-ended questions, were mandatory and required a response. Respondents were required to complete outstanding items before leaving the survey page. To minimise the risk of the participants completing the survey more than once, Qualtrics places a browser cookie on response submission to withhold repeated attempts. Any suspected duplicate submissions were flagged for follow-up post data collection. Additional security measures were taken to ensure the survey was not completed by bots, with a bot detection and security scan feature enabled in the data field. Further precautions were made to block search engines from including the survey in their search results to ensure respondents were from the assigned inclusion criteria. Only non-duplicate and completed responses from eligible participants were included in the analysis dataset.

### 2.5 Patient and public involvement

Patients and members of the public were not involved in the design, conduct, or reporting of this study. The work was delivered within the constraints of an eight-month pilot grant, which limited the feasibility of establishing formal patient and public involvement structures.

### 2.6 Analyses

The data were analysed using descriptive statistics to calculate participant characteristics, frequency and percentiles were employed for categorical variables. Additionally, the Likert scales were analysed and displayed descriptively to align with current themes and study outcomes.

Survey items used Likert⍰type ordinal scales, and the distributions of responses were non⍰normal; hence, a Mann-Whitney Test was conducted to interrogate whether differences existed between caregiver and managerial staff answers in relation to the questions investigating *staff’s and residents’ thermal comfort*. Statistical analyses were carried out using SPSS (version 28.1; Chicago, USA).

For the open-ended questions, thematic analysis was used (40), to examine patterns of meaning within qualitative data (41). It provides a systematic yet flexible approach that supports interpretive analysis, allowing researchers to explore both the frequency and the contextual significance of emerging themes (41). The thematic analysis followed the 6-step framework (42); 1: transcript creation and data familiarisation, 2: keyword identification, 3: code selection, 4: theme development, 5: conceptualisation through interpretation of keywords, codes, and themes. Phase 6—the construction of an overarching conceptual model—was not undertaken, as the breadth and depth of the open-ended responses were not sufficient to support a model of appropriate analytical robustness. An inductive, semantic approach was employed within a realist epistemological framework, meaning that coding was data-driven, themes were based on the explicit content of participants’ responses, and participants’ accounts were treated as reflecting their genuine experiences and perspectives (43). Due to the relatively small qualitative data set, data was manually coded using Excel Software (Microsoft Corporation, Redmond, WA, USA).

To ensure the trustworthiness of the findings, we aligned the analytic procedures with recognised standards of credibility, dependability, confirmability, and transparency in qualitative research (44, 45). Additionally, reflexive practice was undertaken throughout the analytic process, with researchers critically examining how their own assumptions, disciplinary backgrounds, and interpretive positions might shape coding decisions and theme development. Guided by recommendations for methodological rigor (46, 47), researcher HB conducted two rounds of reliability checks followed by an independent review of the coding framework by researcher NKE. Both researchers then met to discuss areas of convergence and divergence, resolving all discrepancies through consensus without the need for a third reviewer or moderator. The same procedure was applied during theme refinement to ensure coherence, credibility, and dependability of the analysis. An audit trail was maintained throughout to support transparency in analytic decision-making.

## 3. Results

### 3.1 Participant demographics

Across the UK and Ireland, 225 care home staff participated in this survey. Participants were distributed across managerial (39%) and front-line caregiver (61%) roles, with respondents from private-, state- and charity-funded homes. Participant data is outlined in Table 1.

**Table 1.** Participant data.

|  |  |  |
| --- | --- | --- |
| *N* |  | 225 |
| Age<br>(years; *Mean ± SD*) |  | 39 ± 10 |
| Gender<br>(*N [%]*) | Female | 151 [67%] |
|  | Male | 72 [32%] |
|  | Prefer not to say | 2 [1%] |
| Job role<br>(*N [%]*) | Managerial | 88 [39%] |
|  | Care Giver | 137 [61%] |
| Years worked in the care sector<br>(*Mean ± SD*) |  | 10 ± 7 |
| Care home type<br>(*N [%]*) | Care home | 75 [33%] |
|  | Nursing home | 86 [38%] |
|  | Dual registered | 56 [25%] |
|  | Other | 8 [4%] |
| Funding source<br>(*N*)* | Private | 165 |
|  | State/Government | 92 |
|  | Charity | 29 |

\*Note: some care home had multiple funding sources and therefore, were included in multiple categories.

### 3.2 Staff and residents’ thermal comfort

No statistically significant differences were found between managerial staff and care givers in response to the questions outlined in this section (*p* > 0.069), hence all staff responses are considered together.

Survey responses within this section indicated that most staff: (*i*) complained of being too hot three or more times per day (148/225; 66%); and (*ii*) reported residents to complain about being too hot on average two times per day (77/225; 34%; Figure 1A). There was consistent agreement amongst responders that extreme heat negatively affected their ability to carry out tasks (203/225; 90%) and that staff perceived these events to impact the thermal comfort and health of the residents (206/225; 92%).

**Figure 1.**
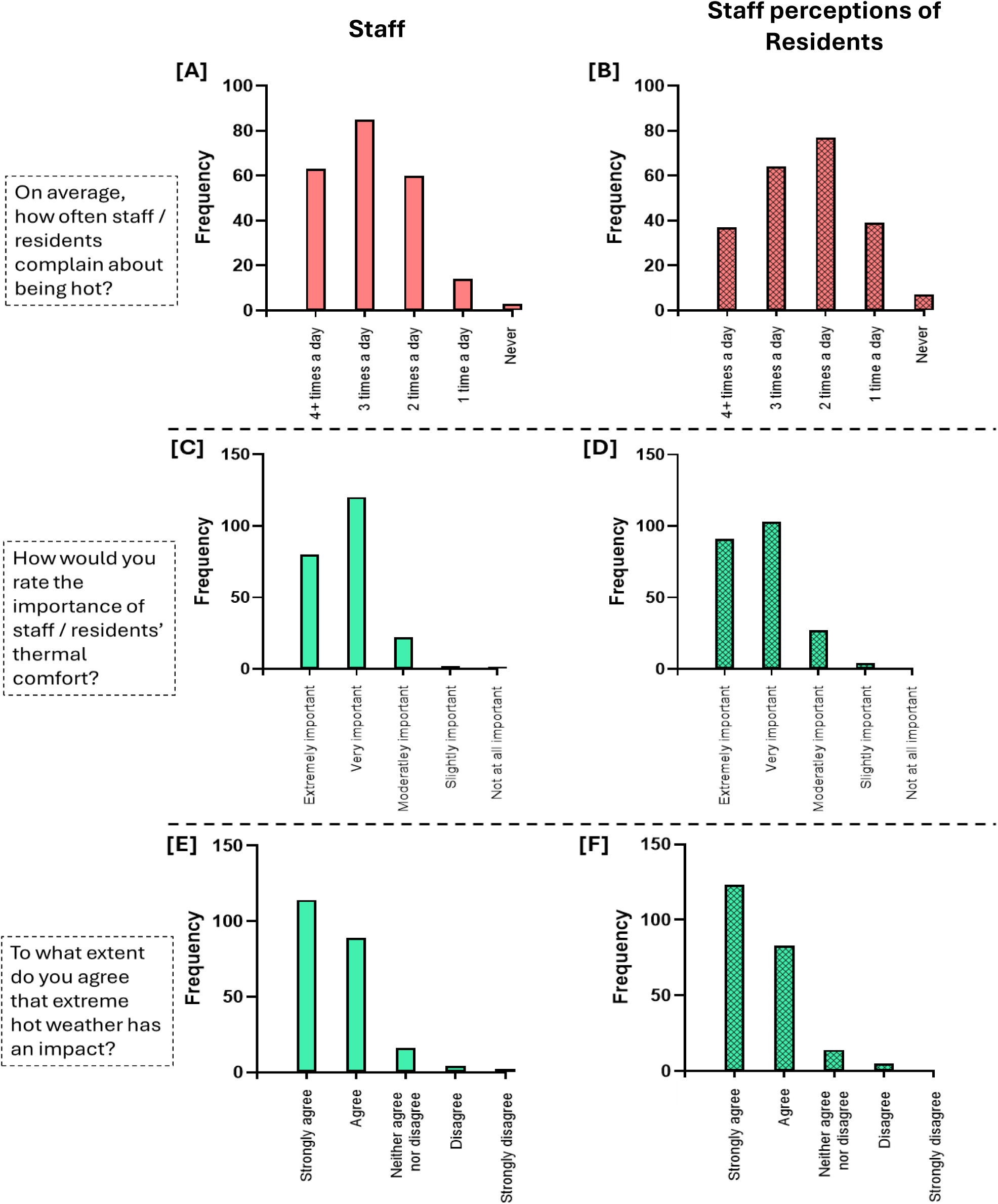
Histograms displaying the frequency by which [A] staff and [B] staff perceive residents to complain of being too hot per day, the staff’s perceived importance of [C] staff and [D] residents’ thermal comfort, and the perceived impact of extreme hot weather events on [E] staff’s ability to perform tasks and [F] residents’ thermal comfort and health.

Survey responses revealed that the majority of staff deemed their own thermal comfort (i.e., 200/225; 89%), as well as the thermal comfort of their residents (i.e., 194/225; 86%), to be very important (Figure 1).

When asked to rate the importance of six day-to-day tasks during extreme hot weather, staff reported medical needs (e.g., medication/turning) to have the greatest importance (i.e., highest median rank = 5), followed by residents’ thermal needs/comfort (e.g., keeping them warm/cool; rank = 4.5), personal care tasks (e.g., dressing and washing residents; rank = 4), meal support (e.g., ensuring they are fed and hydrated; rank = 3), providing company (e.g., chatting; rank = 2) and admin tasks (e.g., recording essential information; rank = 2).

Two thirds (148/225; 66%) of surveyed staff generally reported satisfaction with their thermal environment, with 19% being extremely satisfied and 47% being somewhat satisfied. However, when probed to highlight what caused dissatisfaction with their work-place thermal environment, the most common response was the temperature, followed by air movement (or thereby lack of), and then the clothing provided (Figure 2).

**Figure 2.**
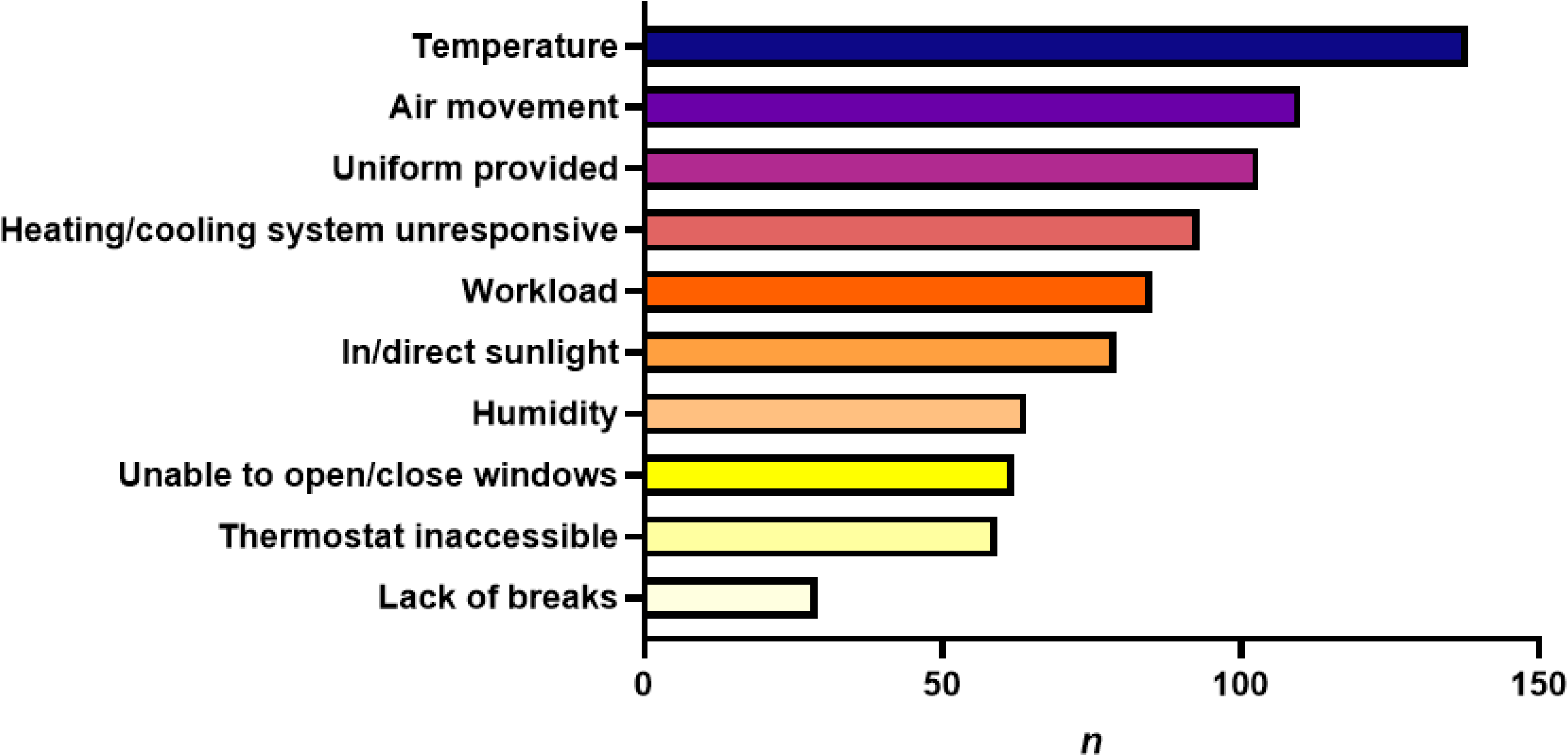
Staff reasons for dissatisfaction with their workplace thermal environment.

### 3.3 Heat resilience strategies—knowledge and implementation

The majority of responders (215/225; 96%) reported maintaining a stable temperature in the summer to be a challenge. Of the surveyed 225 staff, 214 (95%) staff members were aware that older adults are at greater risk during extreme hot weather. Most staff members (i.e., 200/225; 89%) were cognisant of guidance to protect residents during extreme hot, and how to spot the signs of heat illness during extreme heat events, with 197 (87%) staff members assessing individual residents for signs of heat illness at least once per day.

During extreme hot weather events, care home staff reported that opening windows (169/225; 75%) and the use of fans (166/225; 74%) were the most used cooling strategies. Conversely, control of the thermostat (88/225; 39%) and access to air conditioning (108/225; 48%) were least common (Figure 3).

**Figure 3.**
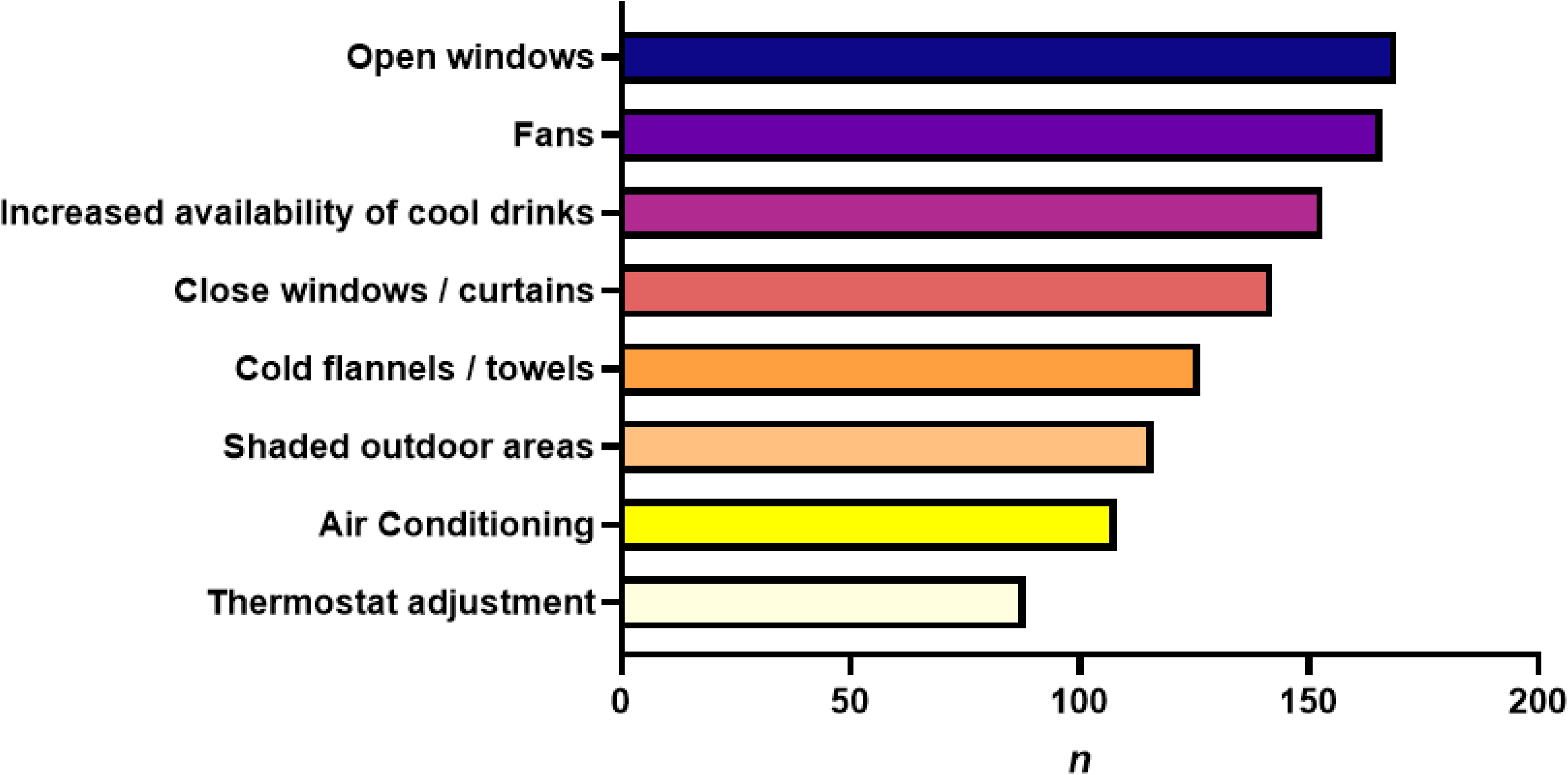
Cooling strategies currently used in care homes to support residents during extreme hot weather.

### 3.4 Barriers to providing heat resilience strategies

Eighty-seven participants (40%) completed the optional open-ended questions regarding the barriers to providing heat resilience strategies. Five key themes emerged regarding the barriers to care home staff providing cooling strategies during extreme hot weather and four key themes arose as potential strategy improvements to manage resident thermal comfort and heat illness (Table 2).

**Table 2.**
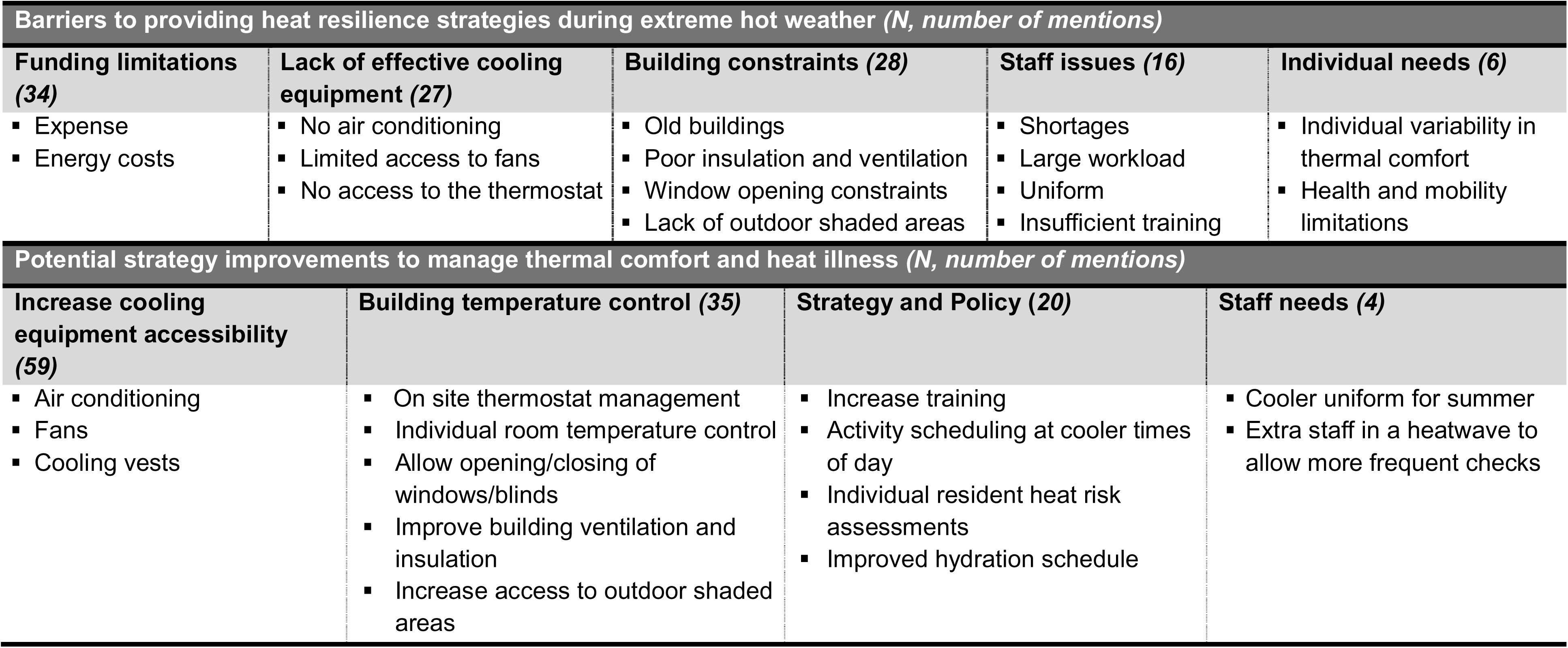
Dominant themes from the thematic analysis assessing the barriers to providing heat resilience strategies.

| **Barriers to providing heat resilience strategies during extreme hot weather (N, number of mentions)** |  |  |  |  |
| --- | --- | --- | --- | --- |
| **Funding limitations (34)** | **Lack of effective cooling equipment (27)** | **Building constraints (28)** | **Staff issues (16)** | **Individual needs (6)** |
| ▪ Expense<br>▪ Energy costs | ▪ No air conditioning<br>▪ Limited access to fans<br>▪ No access to the thermostat | ▪ Old buildings<br>▪ Poor insulation and ventilation<br>▪ Window opening constraints<br>▪ Lack of outdoor shaded areas | ▪ Shortages<br>▪ Large workload<br>▪ Uniform<br>▪ Insufficient training | ▪ Individual variability in thermal comfort<br>▪ Health and mobility limitations |
| **Potential strategy improvements to manage thermal comfort and heat illness (N, number of mentions)** |  |  |  |  |
| **Increase cooling equipment accessibility (59)** | **Building temperature control (35)** | **Strategy and Policy (20)** | **Staff needs (4)** |  |
| ▪ Air conditioning<br>▪ Fans<br>▪ Cooling vests | ▪ On site thermostat management<br>▪ Individual room temperature control<br>▪ Allow opening/closing of windows/blinds<br>▪ Improve building ventilation and insulation<br>▪ Increase access to outdoor shaded areas | ▪ Increase training<br>▪ Activity scheduling at cooler times of day<br>▪ Individual resident heat risk assessments<br>▪ Improved hydration schedule | ▪ Cooler uniform for summer<br>▪ Extra staff in a heatwave to allow more frequent checks |  |

The limitation of funding for cooling equipment and the energy costs to run this were a recurring theme in the survey responses. Several respondents (*N* = 39) mentioned budget constraints more broadly, while other specifically highlighted the issue of:

> *“The expense of installation of cooling systems throughout the home”.*

> *“Air conditioning units are constantly under repair due to their age” causing “expensive maintenance costs” which “without any energy savings in place, i.e., solar panels, is a vast costing to business.”*

The lack of effective cooling equipment was also highlighted as a key barrier to maintaining thermal comfort during extreme hot weather:

> *“A lack of air conditioning means the top floors are ovens during the summer” “Fans just blow hot air around”*

This barrier of limited cooling equipment was further emphasised by the need for increased accessibility to cooling equipment being a key theme for potential improvement strategies. Many respondents (*N* = 27) spoke of the need for permanent air conditioning installation, at least in key rooms, such as the communal lounges and dining rooms.

Another key theme that appeared as a barrier to cooling, as well as being highlighted as an area for improvement, was that of building constraints. An inability to open/close windows to increase ventilation, old buildings with poor insulation and a lack of access to shaded outdoor spaces were mentioned regularly in the survey responses (*N* = 28). Some respondents mentioned the need to:

> *“Review and upgrade insulation to keep indoor temperatures stable”*

Whilst others highlighted issues of the building and care home constraints:

> *“Poor building layout, especially in older facilities” and “Difficulty in maintaining consistent indoor temperatures in older or larger buildings”*

> *“For safety, all windows have restrictors so difficult to get airflow in bedrooms”*

The thematic analysis also highlighted staff issues and individual resident needs as a barrier to cooling strategies. These themes depicted the issues of staff shortages and excessive workload as well as insufficient staff training to deal with extreme heat periods:

> *“Insufficient staff to monitor residents frequently during heatwaves” and a “high staff workload makes it difficult to implement extra cooling measures”.*

The added complication of individual resident needs acts as a further barrier whereby:

> *“Some residents mobility or health conditions can make it difficult to implement all cooling strategies consistently” and “residents sometimes have sensory/personal care needs that stop staff from being able to give cold flannels or moving to a cooler room due to mobility issues/health”.*

These barriers feed into the potential improvement themes of updating strategy and policy as well as staffing needs, with the highlighted requirements for increased staff training, individual heat risk assessments for residents and staff, and increased staff numbers during heatwaves:

> *“I’d add continuous temperature monitoring, individual heat-risk profiles, and proactive cooling measures to make thermal comfort management more preventative.”*

> We need *“a way to cool staff who are hot through movement whilst keeping residents who are cold through being sedentary warm”* and to *“create individual heat plans for residents with high vulnerability”*.

## 4. Discussion

The aim of this study was to understand the impact of heatwaves on UK and Irish care home staff’s and staff’s perception of residents’ health and comfort, evaluating the heat resilience strategies employed by care home staff during extreme heat events. The cross-sectional survey also explored the barriers and facilitators to implementing heat resilience strategies. Leveraging a UK and Ireland-wide recruitment of care home staff’s experiences, challenges, and adaptation strategies during heatwaves, we were able to collect insights on heat (mal-)adaptation from a diverse set of care home staff (*N* = 225). The results highlighted several key themes, notably that care home staff:

*i)* perceive extreme hot weather events (i.e., heatwaves) to negatively impact their ability to work and the residents’ thermal comfort and health;
*ii)* have an understanding of the risk of extreme heat on residents and knowledge of how to monitor heat illnesses;
*iii)* face several barriers to implement effective heat resilience strategies to ensure a well maintained and safe thermal environment for themselves and residents.

### 4.1. Staff and residents’ health, comfort and vulnerability

The findings of this study reinforce the growing recognition that care home staff are themselves vulnerable to extreme heat exposure, and that thermal risk in care settings extends far beyond traditionally defined “at-risk” groups (19, 20, 48). While older residents remain highly susceptible to heat-related illness due to age-related impairments in thermoregulation (12–14), our data show that staff also frequently experience thermal discomfort, with two-thirds reporting feeling too hot multiple times per day. The spread of data shown in Figure 1 demonstrates that staff perceive themselves to complain of over-heating more regularly than they perceive the residents to, which aligns to recent findings that care home residents tend to be more tolerant of extreme temperature variations than staff (37).

Importantly, the results of this study demonstrated that surveyed staff had good knowledge of the increased vulnerability of older people to heatwaves and of how to assess signs of heat illness—an improvement on the more limited care home staff awareness reported in earlier studies (26, 27). This may reflect improvements in, and increased accessibility of, information sources for social care practitioners following the extreme high temperatures experienced in the UK during the summer of 2022 (17), as these earlier studies pre-dated this period. This suggests that knowledge-based preparedness among frontline staff may be increasing in response to more frequent and memorable heat events.

Care home staff perceived the maintenance of thermal comfort to be of high importance for themselves and the residents, which aligns with previous findings during cold periods in winter (36). Furthermore, care home staff reported that extreme hot weather events negatively impacted both their ability to perform their daily tasks, and residents’ comfort and health (Figure 2). This aligns with recent findings by UKHSA (17) who identified social care practitioners experience occupational and cognitive strain during heatwaves, strain that may subsequently affect the quality and consistency of care delivered.

### 4.2. Heat resilience strategies and barriers

Survey responses revealed that the most frequently used heat-resilience strategies were opening windows (75%) and the use of fans (74%). These approaches are likely widespread because they are low-cost, readily accessible, and require minimal organisational approval or infrastructure. However, the qualitative findings emphasise that both strategies offer only limited relief under the conditions encountered during heatwaves. Staff frequently noted that window opening was restricted for safety reasons—a long-standing regulatory requirement in UK and Irish care homes (22)—making it difficult to achieve adequate ventilation during periods of extreme heat. As one participant explained, “for safety, all windows have restrictors so [it is] difficult to get airflow in bedrooms”. Similarly, staff described the limited effectiveness of fans in overheated indoor environments, with several respondents stating that “fans just blow hot air around”. These reports echo previous suggestions that natural ventilation (49) and mechanical air movement through the use of fans (50, 51) can have detrimental effects on cooling when indoor temperatures already exceed recommended thresholds.

A dominant barrier to implementing more effective cooling measures related to funding limitations (*N* = 34) and inadequate or ageing cooling equipment (*N* = 27). Staff frequently commented on the prohibitive costs associated with installing or maintaining air-conditioning systems, such as “the expense of installation of cooling systems throughout the home” and “air conditioning units constantly under repair due to their age,” the latter of which imposed “expensive maintenance costs… without any energy savings in place”. These insights highlight the financial vulnerability of a sector already under pressure (52), and help explain the continued reliance on low-cost but suboptimal adaptations.

In addition to equipment-related constraints, staff highlighted substantial structural limitations within the buildings themselves (*N* = 28). Many care homes, particularly older facilities, were described as having poor insulation, inadequate ventilation, and layouts that made it difficult to maintain stable and comfortable indoor temperatures. Participants noted issues such as “poor building layout, especially in older facilities” and “difficulty in maintaining consistent indoor temperatures in older or larger buildings”. These concerns align with recent research demonstrating that older care homes with inadequate insulation are at heightened risk of summertime overheating and reduced indoor air quality (53). Yet, as with cooling technologies, upgrading the building fabric or infrastructure would be severely constrained by limited budgets.

Finally, staffing challenges emerged as an important barrier to effective heat-resilience implementation (*N* = 16). Staff shortages and increased workloads were repeatedly described as limiting the ability to monitor residents or put additional cooling measures in place during heatwaves. One respondent noted that the “high staff workload makes it difficult to implement extra cooling measures,” while another explained that “insufficient staff [are available] to monitor residents frequently during heatwaves”. These pressures not only reduce the capacity to support residents but may also increase the risk to staff themselves, who must carry out physically and cognitively demanding work in already overheated environments.

### 4.3. Areas for strategic improvement

This study highlights several strategies that care home staff believe would strengthen heat resilience and support both resident and staff safety. The most suggested improvement was increasing access to effective cooling equipment (*N* = 59). However, in line with the financial pressures already being experienced by this sector (52), the feasibility of installing or upgrading air-conditioning systems is likely limited. The second most frequently proposed strategy (*N* = 35) related to improving local control of building temperatures. By enabling on-site teams to adjust thermostats or heating systems, this would allow more proactive management of indoor conditions; thus, providing a straightforward, low-cost approach to improve heat resilience. Similarly, respondents suggested increasing flexibility in building ventilation, such as allowing wider opening of windows; although this would require careful review of existing safety regulations to ensure resident protection is maintained.

Beyond equipment and environmental controls, staff also identified a set of organisational and staffing-related strategies (*N* = 20) that could offer low-cost, high-impact improvements. While participants already demonstrated strong awareness of heat-related risks, they noted that additional training could be beneficial. For example, further training could help to optimise the use and timing of low-cost cooling measures. Staff also proposed rescheduling activities to cooler times of day, hence reducing heat strain on both residents and workers. Although increasing staffing numbers during heatwaves was suggested by the participants, it must be acknowledged that this is unlikely to be achievable within current workforce and budget constraints. A more feasible alternative may be the introduction of modified shift patterns that incorporate regular rest or cooling breaks, helping to mitigate physical and cognitive fatigue during prolonged heat exposure (19, 20). Such adjustments could form a practical component of improved heatwave-resilience policies.

Together, these recommendations point to a need for combined investment in environmental controls alongside flexible operational and staffing approaches, many of which are low-cost and immediately implementable within existing constraints.

### 4.4. Limitations

This study has several limitations. First, survey responses were collected between March and September 2025, which may introduce recall bias, particularly for respondents completing the survey in spring and who may have reflected on heatwaves from 2024. To mitigate over!Zlinterpretation, we emphasise that primary outcomes focused on knowledge, perceptions, and reported practices (e.g., awareness of heat risks, signs of heat illness, and commonly used strategies), which are comparatively stable over time; nevertheless, perceptions of specific events and their intensity may be affected by recall.

Second, findings may not be fully generalisable beyond the UK and Irish social care context. Care homes differ internationally in building stock, regulation, staffing models, and access to cooling technologies; even within the UK and Ireland, variability by home type, size, funding model, and building age may limit external validity. While many challenges identified here (e.g., ageing buildings, restricted ventilation, funding constraints) are likely shared across settings experiencing more frequent heatwaves, transferability should be inferred cautiously and confirmed by local evaluation.

## 5. Conclusion

Extreme heat continues to pose a growing risk to the health and wellbeing of older people in care homes; yet, as importantly highlighted in this study, extreme heat also can be a risk to the staff who support them. While staff demonstrated strong awareness of heat!Zlrelated risks and how to recognise heat illness, their ability to protect residents is limited by inadequate cooling equipment, ageing buildings, restricted ventilation, staffing pressures, and financial constraints. These findings highlight that current thermal!Zlresilience measures are insufficient and often reactive. Practical, low!Zlcost improvements—such as enhanced temperature control, greater access to basic cooling devices, targeted training, and operational adjustments during extreme heat—could better safeguard both staff and residents. By capturing the lived experiences of frontline care workers across diverse settings, this study provides evidence urgently needed to inform public health policy, strengthen heatwave preparedness guidance, and support more resilient, equitable, and sustainable care home environments as extreme heat events become increasingly common.

## Data availability

Data will be made available upon publication at the University of Southampton data repository (PURE; URL to be activated upon publication).

## Author contributions

HB, JW, DF and NKE conceived and designed research. HB, JW, PRW, DF and NKE recruited and conducted the data collection. HB and JW analysed data. HB, JW, DF and NKE interpreted results. HB prepared figures. HB, JW and NKE drafted manuscript. HB, JW, PABJ, PRW, DF and NKE edited and revised the manuscript. HB, JW, PABJ, PRW, DF and NKE approved final version of the manuscript.

## Acknowledgements

The authors would like to thank all the volunteers who partook in the survey. For the purpose of open access, the author has applied a Creative Commons attribution license (CC BY) to any Author Accepted Manuscript version arising from this submission.

## References

1. Lowe J, Bernie D, Bett PE, Bricheno LM, Brown S, Calvert D, et al., editors. UKCP 18 Science Overview Report November 2018 (Updated March 2019 )2019: MetOffice.

2. Ebi KL, Vanos J, Baldwin JW, Bell JE, Hondula DM, Errett NA, et al. Extreme Weather and Climate Change: Population Health and Health System Implications. Annu Rev Public Health. 2021;42:293–315.

3. Ebi KL, Capon A, Berry P, Broderick C, de Dear R, Havenith G, et al. Hot weather and heat extremes: health risks. Lancet. 2021;398(10301):698–708.

4. Ballester J, Quijal-Zamorano M, Méndez Turrubiates RF, Pegenaute F, Herrmann FR, Robine JM, et al. Heat-related mortality in Europe during the summer of 2022. Nat Med. 2023;29(7):1857–66.

5. Ons U. Excess mortality during heat-periods: 1 June to 31 August 2022. In: Ons U, editor.: GOV.UK; 2022.

6. Addison TEJ, Blake DM, Coleman P, Lock F, Loud E, Mackenzie A, et al. Physiological considerations for maximum indoor temperatures. PLOS Climate. 2025;4(6):e0000642.

7. WHO WHO. Heat claims more than 175 000 lives annually in the WHO European Region, with numbers set to soar. In: Europe W, editor. WHO2024.

8. Ferrari AU, Radaelli A, Centola M. Invited review: aging and the cardiovascular system. J Appl Physiol (1985). 2003;95(6):2591–7.

9. Schmidt MD, Notley SR, Meade RD, Akerman AP, Rutherford MM, Kenny GP. Revisiting regional variation in the age-related reduction in sweat rate during passive heat stress. Physiol Rep. 2022;10(7):e15250.

10. Meade RD, Akerman AP, Notley SR, McGinn R, Poirier P, Gosselin P, Kenny GP. Physiological factors characterizing heat-vulnerable older adults: a narrative review. Environ Int. 2020;144:105909.

11. Guergova S, Dufour A. Thermal sensitivity in the elderly: a review. Ageing Res Rev. 2011;10(1):80–92.

12. Kovats RS, Hajat S. Heat Stress and Public Health: A Critical Review. Annual Review of Public Health. 2008;29(Volume 29, 2008):41–55.

13. Sheridan SC, Kalkstein AJ, Kalkstein LS. Trends in heat-related mortality in the United States, 1975–2004. Natural Hazards. 2009;50(1):145–60.

14. Kyselý J, Kříž B. Decreased impacts of the 2003 heat waves on mortality in the Czech Republic: an improved response? International Journal of Biometeorology. 2008;52(8):733–45.

15. Brown S, and Walker G. Understanding heat wave vulnerability in nursing and residential homes. Building Research & Information. 2008;36(4):363–72.

16. Gupta R, and Gregg M. Care provision fit for a warming climate. Architectural Science Review. 2017;60(4):275–85.

17. UKHSA. Research exploring the experience of social care practitioners in relation to extreme temperatures. In: UK Health Security Agency GU, editor.: GOV.UK; 2024.

18. Committee EA. Heat resilience and sustainable cooling – Report Summary. In: Parliament U, editor. 2024.

19. Wibowo R, Satow M, Quartucci C, Weinmann T, Koller D, Daanen HAM, et al. Impact of heat stress and protective clothing on healthcare workers: health, performance, and well-being in hospital settings. Annals of Work Exposures and Health. 2025;69(6):665–75.

20. Davey SL, Lee BJ, Robbins T, Thake CD. Prevalence of occupational heat stress across the seasons and its management amongst healthcare professionals in the UK. Applied Ergonomics. 2024;118:104281.

21. Meerow S, Keith L. Planning for Extreme Heat. Journal of the American Planning Association. 2022;88(3):319–34.

22. Gupta R, Howard A, Davies M, Mavrogianni A, Tsoulou I, Oikonomou E, Wilkinson P. Examining the magnitude and perception of summertime overheating in London care homes. Building Services Engineering Research & Technology. 2021;42(6):653–75.

23. Forcada N, Gangolells M, Casals M, Tejedor B, Macarulla M, Gaspar K. Summer thermal comfort in nursing homes in the Mediterranean climate. Energy and Buildings. 2020;229:110442.

24. Smythe A, Jenkins C, Galant-Miecznikowska M, Bentham P, Oyebode J. A qualitative study investigating training requirements of nurses working with people with dementia in nursing homes. Nurse Education Today. 2017;50:119–23.

25. Cowan DT, Roberts JD, Fitzpatrick JM, While AE, Baldwin J. Nutritional status of older people in long term care settings: current status and future directions. International Journal of Nursing Studies. 2004;41(3):225–37.

26. Greene C, Canning D, Wilson J, Bak A, Tingle A, Tsiami A, Loveday H. I-Hydrate training intervention for staff working in a care home setting: An observational study. Nurse Education Today. 2018;68:61–5.

27. Gupta R, Barnfield L, Gregg M. Overheating in care settings: magnitude, causes, preparedness and remedies. Building Research & Information. 2017;45(1-2):83–101.

28. CareUK. What’s the difference between a care home and a nursing home? 2025 [02/02/2026]. Available from: https://www.careuk.com/help-advice/what-s-the-difference-between-a-care-home-and-a-nursing-home#:~:text=Wondering%20if%20residential%20care%20or,tasks%20like%20cleaning%20and%20cooking.

29. yoon Yi C, Childs C, Peng C, Robinson D. Thermal comfort modelling of older people living in care homes: An evaluation of heat balance, adaptive comfort, and thermographic methods. Building and Environment. 2022;207:108550.

30. Yu J, Hassan MT, Bai Y, An N, Tam VW. A pilot study monitoring the thermal comfort of the elderly living in nursing homes in Hefei, China, using wireless sensor networks, site measurements and a survey. Indoor Built Environ. 2020;29(3):449–64.

31. van Hoof J, Schellen L, Soebarto V, Wong J, Kazak J. Ten questions concerning thermal comfort and ageing. Building and Environment. 2017;120:123–33.

32. Hughes C, Natarajan S. Summer thermal comfort and overheating in the elderly. Building Services Engineering Research and Technology. 2019;40(4):426–45.

33. Tsoulou I. Indoor Overheating, Climate Resilience, and Adaptation of Care Settings. The Palgrave Handbook of Climate Resilient Societies. 2021.

34. Black D, Veitch C, Wilson L. Heat-Ready: heatwave awareness, preparedness and adaptive capacity in aged care facilities. 2013.

35. Jiao Y, Yu Y, Yu H, Wang F. The impact of thermal environment of transition spaces in elderly-care buildings on thermal adaptation and thermal behavior of the elderly. Building and Environment. 2023;228:109871.

36. Walker G, Brown S, Neven L. Thermal comfort in care homes: vulnerability, responsibility and ‘thermal care’. Building Research & Information. 2016;44(2):135–46.

37. Tartarini F, Cooper P, Fleming R. Thermal perceptions, preferences and adaptive behaviours of occupants of nursing homes. Building and Environment. 2018;132:57–69.

38. Okwuofu-Thomas B. Analysis of heatwave response plans and adaptation to cope with heatwaves now and in the future in aged care facilities: Macquarie University, Faculty of Science and Engineering, Department of …; 2017.

39. Boyson C, Taylor S, Page L. The national heatwave plan–a brief evaluation of issues for frontline health staff. PLoS currents. 2014;6:ecurrents. dis. aa63b5ff4cdaf47f1dc6bf44921afe93.

40. Braun V, Clarke V. Reflecting on reflexive thematic analysis. Qualitative Research in Sport, Exercise and Health. 2019;11(4):589–97.

41. Alhojailan MI, Ibrahim M. Thematic analysis: A critical review of its process and evaluation. West east journal of social sciences. 2012;1(1):39–47.

42. Naeem M, Ozuem W, Howell K, Ranfagni S. A Step-by-Step Process of Thematic Analysis to Develop a Conceptual Model in Qualitative Research. International Journal of Qualitative Methods. 2023;22:16094069231205789.

43. Ahmed SK, Mohammed RA, Nashwan AJ, Ibrahim RH, Abdalla AQ, Ameen BMM, Khdhir RM. Using thematic analysis in qualitative research. Journal of Medicine, Surgery, and Public Health. 2025;6:100198.

44. Haq ZU, Rasheed R, Rashid A, Akhter S. Criteria for assessing and ensuring the trustworthiness in qualitative research. International Journal of Business Reflections. 2023;4(2).

45. Cope DG, editor Methods and meanings: credibility and trustworthiness of qualitative research. Oncology nursing forum; 2014.

46. Hruschka DJ, Schwartz D, St.John DC, Picone-Decaro E, Jenkins RA, Carey JW. Reliability in Coding Open-Ended Data: Lessons Learned from HIV Behavioral Research. Field Methods. 2004;16(3):307–31.

47. Belotto MJ. Data analysis methods for qualitative research: Managing the challenges of coding, interrater reliability, and thematic analysis. The qualitative report. 2018;23(11):2622–33.

48. Filingeri D, Koch Esteves N. How hot is too hot for people? A review of empirical models of perceptual, physiological and functional limits of human heat tolerance. Experimental Physiology. 2025.

49. UKHSA. Beat the heat: staying safe in hot weather. In: UK Health Security Agency GU, editor.: GOV.UK; 2025.

50. Morris NB, Chaseling GK, English T, Gruss F, Maideen MFB, Capon A, Jay O. Electric fan use for cooling during hot weather: a biophysical modelling study. Lancet Planet Health. 2021;5(6):e368–e77.

51. Meade RD, Notley SR, Kirby NV, Kenny GP. A critical review of the effectiveness of electric fans as a personal cooling intervention in hot weather and heatwaves. The Lancet Planetary Health. 2024;8(4):e256–e69.

52. Cousins C, Burrows R, Cousins G, Dunlop E, Mitchell G. An overview of the challenges facing care homes in the UK. Nursing Older People. 2016;28(9).

53. Tsoulou I, Jain N, Oikonomou E, Petrou G, Mavrogianni A, Gupta R, et al. Indoor environmental quality trade-offs due to summertime natural ventilation in London care homes. Journal of Physics: Conference Series. 2023;2600(9):092013.

