## supplementary material for "Understanding the Impact of Heatwaves on UK Care Homes: A National Survey of Staff Experiences, Challenges, and Adaptation Strategies"

ThermoCare Survey

Start of Block: Information and Consent

**Survey Information**

The purpose of this online survey is to gain insights into the impact of extreme hot weather events on **elderly** care home staff and residents' health, comfort and vulnerability. With this knowledge, we hope to guide policy to improve staff and elderly resident comfort and minimise heat-related issues, particularly with the increasing frequency of heatwaves.

An information sheet further detailing what the research is about and what is required of you can be found here: Participant information sheet

**Inclusion criteria:**

- Aged 18 or over.
- Employed staff working in an **elderly** care home / nursing home.
- Holds either a Managerial (e.g. Manager, Deputy Manager, Team Leader, Administration) or Care Giver (e.g. Nurse, Care worker, Senior Care Worker, Personal Assistant) role.

**Exclusion criteria:**

- Unable to complete the survey in English.
- Residents of the care home.
- Non-Care Giver/ Managerial roles (e.g. Catering, Maintenance, Transport, Housekeeping, Activity Coordinator).

End of Block: Information and Consent

Start of Block: Consent

Q2 I have read and understood the attached information sheet and meet the required inclusion criteria to participate in this survey.

- I consent to participating in this survey.
- I agree for my data to be used for the purpose of the study.
- I am aware all data will be anonymous.
- I understand my participation is voluntary and that I can withdraw at any time for any reason without my participation rights being affected.
- I am 18 years or older.
- Yes (1)
- No (2)

Skip To: End of Survey If Q2 = No

Q3 During this survey there might be some terms you are unfamiliar with, here we have provided you with some definitions to help explain.

- ***Extreme hot weather:*** *When thinking about extreme hot weather, please refer to days when you have experienced temperatures outside of the normal. These days might have been highlighted during weather reports on the news or from your local Met Office using a coding system such as yellow, amber/orange or red weather warnings.*
- ***Thermal comfort:*** This is *a feeling of satisfaction with the current state of the indoor or outdoor temperature environment, with the absence of desire for change.*
- ***Cooling strategies:*** *A method or approach to regulate the temperature of a room or person during periods of extreme hot weather.*
- ***Heat illness:*** *A range of disorders caused by an increased body temperature through changes in environmental conditions or physical exertion.*

End of Block: Consent

Start of Block: Demographic Questions

Q4 How old are you?

- Years (1) __________________________________________________

Q5 What is your gender?

- Male (1)
- Female (2)
- Non-binary / third gender (3)
- Prefer not to say (4)
- Other (5)

Q6 What is your role in the care home?

- Managerial (e.g Manager, Deputy Manager, Team Leader, Administration) (1)
- Care Giver (e.g Nurse, Care worker, Senior Care Worker, Personal Assistant) (2)

Q7 What is your job title?

________________________________________________________________

Q8 How long have you worked in your current role? Please state to the closest year.

- Years (1) __________________________________________________

Q9 How long have you worked at the present care home? Please state to the closest year.

- Years (1) __________________________________________________

Q10 How long have you worked in the care home sector? Please state to the closest year.

- Years (1) __________________________________________________

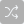

Q11 What country is the care home based in?

- United Kingdom (1)
- Ireland (2)
- Australia (3)
- New Zealand (4)
- South Africa (5)
- United States of America (6)
- Canada (7)
- Singapore (9)
- Other (10) __________________________________________________

Q12 Please write the city and state/region that the care home is based in. E.g. Southampton, Hampshire

________________________________________________________________

Q13 How many residents are currently living in the care home (approximate)?

- Number (1) __________________________________________________

Q14 Is the care home funded by the state/government or private channels? Select all that apply.

- State/Government (1)
- Private (2)
- Charity (3)

Q15 What type of care home is it? **Care home:** helps with personal care such as washing, dressing, taking medication and going to the toilet **Nursing home:** offers personal care as well as 24-hour assistance from qualified nurses **Dual registered care home**: accepts residents who need both personal care and nursing care, i.e. if someones needs increase over time, they won't have to move to a different home.

- Care home (1)
- Nursing home (2)
- Dual registered care home (3)
- Other (4) __________________________________________________

End of Block: Demographic Questions

Start of Block: Main Survey - Staff Knowledge

Q16 Do you think elderly people more likely to face health risks during extreme hot weather?

- Yes (1)
- No (2)
- Unsure (3)

Q17 Are you aware of any guidance regarding safe temperatures for care home residents during extreme hot weather?

- Yes (1)
- No (2)
- Unsure (3)

Q18 Do you have training on how to appropriately monitor and protect residents during extreme hot weather?

- Yes (1)
- No (2)
- Unsure (3)
- Currently under development (4)
- Other (5) __________________________________________________

Q19 Has your strategy and guidance regarding extreme hot weather and resident safety changed over the years?

- Yes (if so, when was the latest change [approximate]) (1) __________________________________________________
- No (2)
- Unsure (3)

Q20 How capable are the residents to independently manage their thermal comfort?

- Very capable (1)
- Capable (2)
- Neither capable, nor uncapable (3)
- Uncapable (4)
- Very uncapable (5)

End of Block: Main Survey - Staff Knowledge

Start of Block: Main Survey - Strategy

Q21 How regularly are indoor temperatures monitored in the care home?

- Very often (4+ times per day) (1)
- Often (1-3 times per day) (2)
- Occasionally (more than once per week) (3)
- Rarely (less than once per week) (4)
- Never (5)
- Unsure (6)

Q22 What temperature is the care home central heating/cooling set to in the **summer**? Note Celsius/Fahrenheit

________________________________________________________________

Q23 What temperature is the care home central heating/cooling set to in **winter**? Note Celsius/Fahrenheit

________________________________________________________________

Q24 Who has access to control indoor temperatures in the care home? Select all that apply

- Care givers (1)
- Managerial staff (2)
- Residents (3)
- Visitors (4)
- Other (5) __________________________________________________

Q25 Does your facility have extreme hot weather policy in place?

- Yes (1)
- No (2)
- Unsure (3)
- Currently under development (4)
- Other (5) __________________________________________________

Q26 What cooling strategies, if any, are currently utilised by the care home to support the residents during extreme hot weather? Select all that apply

- None (1)
- Increased availability of cool drinks (2)
- Air conditioning (3)
- Ability to open windows (4)
- Ability to close windows/curtains (5)
- Fans (6)
- Cold flannels / towels (7)
- Shaded outdoor areas (8)
- Thermostat adjustment (9)
- Other (10) __________________________________________________

Q27 During extreme hot weather, how often would you assess individual residents for risk of heat related illnesses?

- Very often (4+ times per day) (1)
- Often (1-3 times per day) (2)
- Occasionally (more than once per week) (3)
- Rarely (less than once per week) (4)
- Never (5)

End of Block: Main Survey - Strategy

Start of Block: Main Survey - Implementation

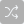

Q28A Please rank these day-to-day tasks in order of importance during extreme hot weather.

______ Personal care tasks (e.g. dressing / washing residents) (1)

______ Providing company (e.g. chatting) (2)

______ Resident thermal needs/comfort (e.g. keeping them warm / cool) (3)

______ Medical needs (e.g. medication / turning) (4)

______ Admin tasks (e.g. recording essential information) (5)

______ Meal support (e.g. ensuring they are fed / hydrated) (6)

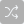

Q28B Please rank these day-to-day tasks in order of importance during extreme hot weather.

______ Personal care tasks (e.g. dressing / washing residents) (1)

______ Providing company (e.g. chatting) (2)

______ Resident thermal needs/comfort (e.g. keeping them warm / cool) (3)

______ Medical needs (e.g. medication / turning) (4)

______ Admin tasks (e.g. recording essential information) (5)

______ Meal support (e.g. ensuring they are fed / hydrated) (6)

Q29 How challenging is maintaining the care home at a stable temperature in the **winter**?

- Extremely challenging (1)
- Very challenging (2)
- Moderately challenging (3)
- Slightly challenging (4)
- Not challenging at all (5)

Q30 How challenging is maintaining the care home at a stable temperature in the **summer**?

- Extremely challenging (1)
- Very challenging (2)
- Moderately challenging (3)
- Slightly challenging (4)
- Not challenging at all (5)

Q31 Who is responsible for looking after the thermal care needs of the residents?  Please select all that apply.

- Myself (1)
- Someone else *please state the role* (2) __________________________________________________
- Themselves (3)

Q32 If someone is displaying signs of heat illness do you know how to spot them?

- Yes (1)
- No (2)
- Unsure (3)

End of Block: Main Survey - Implementation

Start of Block: Main Survey - Resident Comfort

Q33 On average, how often per day do care home **residents** complain about being cold?

- Very often (4+ times a day) (1)
- Often (3 times a day) (2)
- Occasionally (2 times a day) (3)
- Rarely (once a day) (4)
- Never (5)

Q34 On average, how often per day do care home **residents** complain about being hot?

- Very often (4+ times a day) (1)
- Often (3 times a day) (2)
- Occasionally (2 times a day (3)
- Rarely (once a day) (4)
- Never (5)

Q35 How would you rate the importance of **residents'** thermal comfort?

- Extremely important (1)
- Very important (2)
- Moderately important (3)
- Slightly important (4)
- Not at all important (5)

Q36 To what extent do you agree that the thermal environment is impacting the comfort and health of **residents**?

- Strongly agree (1)
- Agree (2)
- Neither agree nor disagree (3)
- Disagree (4)
- Strongly disagree (5)

End of Block: Main Survey - Resident Comfort

Start of Block: Main Survey - Staff Comfort

Q37 On average, how often per day do care home **staff** complain about being cold?

- Very often (4+ times a day) (1)
- Often (3 times a day) (2)
- Occasionally (2 times a day) (3)
- Rarely (once a day) (4)
- Never (5)

Q38 On average, how often per day do care home **staff** complain about being hot?

- Very often (4+ times a day) (1)
- Often (3 times a day) (2)
- Occasionally (2 times a day) (3)
- Rarely (once a day) (4)
- Never (5)

Q39 To what extent do you agree that extreme hot weather impacts your ability to perform daily tasks.

- Strongly agree (1)
- Somewhat agree (2)
- Neither agree nor disagree (3)
- Somewhat disagree (4)
- Strongly disagree (5)

Q40 How would you rate the importance of **staff** members' thermal comfort?

- Extremely important (1)
- Very important (2)
- Moderately important (3)
- Slightly important (4)
- Not at all important (5)

Q41 How satisfied are you with the temperature of your workspace during extreme hot weather?

- Extremely satisfied (1)
- Somewhat satisfied (2)
- Neither satisfied nor dissatisfied (3)
- Somewhat dissatisfied (4)
- Extremely dissatisfied (5)

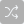

Q42 If you have indicated any dissatisfaction with the temperature in your workspace during extreme hot weather. Please select the source of the discomfort.  Please select all that apply.

- Humidity (high / low) (1)
- Air movement (high / low) (2)
- Temperature (high / low) (3)
- In / direct sunlight (4)
- Clothing provided (5)
- Unable to open or close windows (6)
- Heating / cooling system unresponsive (7)
- Thermostat inaccessible / controlled by others (8)
- Work rate (9)
- Lack of breaks (10)
- None of the above (11)
- Other (13) __________________________________________________

End of Block: Main Survey - Staff Comfort

Start of Block: Main Survey - Open Questions

Q43 Would you like to answer 2 additional open-ended questions?

- Yes (1)
- No (2)

Skip To: End of Block If Q43 = No

Q44 Is there anything you would change to the care home strategy to manage thermal comfort and heat illness?

________________________________________________________________

________________________________________________________________

________________________________________________________________

________________________________________________________________

________________________________________________________________

Q45 Are there any barriers to providing cooling strategies during extreme hot weather?

________________________________________________________________

________________________________________________________________

________________________________________________________________

________________________________________________________________

________________________________________________________________

End of Block: Main Survey - Open Questions

Start of Block: End of Survey

Q46 Would you like to be entered into the prize pot to win a voucher for taking part in this survey? PASSWORD: ThermoCare An email address will need to be provided. This will remain anonymous and it will not be linked to your answers.  Prizes will be drawn 1st September 2025

- Yes (1)
- No (2)

Q47 I am happy to be contacted in the future if I meet the inclusion criteria for upcoming studies conducted by the Skin Sensing Research Group.

- Yes (1)
- No (2)

End of Block: End of Survey
